# Evaluating Life Simple Seven’s influence on brain health outcomes: The intersection of lifestyle and dementia

**DOI:** 10.1101/2024.07.29.24311179

**Authors:** Diandra N Denier-Fields, Ronald E Gangnon, Leonardo A Rivera-Rivera, Tobey J Betthauser, Barbara B Bendlin, Sterling C Johnson, Corinne D Engelman

## Abstract

**Background:** Lifestyle factors have been studied for dementia risk, but few have comprehensively assessed both Alzheimer’s disease (AD) and cerebrovascular disease (CBVD) pathologies.

**Objective:** Our research aims to determine the relationships between lifestyle and various dementia pathologies, challenging conventional research paradigms.

**Methods:** Analyzing 1231 Wisconsin Registry for Alzheimer’s Prevention (WRAP) study participants, we focused on Life Simple Seven (LS7) score calculations from questionnaire data and clinical vitals. We assessed brain health indicators including CBVD, AD, and cognition.

**Results:** Higher (healthier) LS7 scores were associated with better CBVD outcomes, including lower percent white matter hyperintensities and higher cerebral blood flow, and higher Preclinical Alzheimer’s Composite 3 and Delayed Recall scores. No significant associations were observed between LS7 scores and AD markers of amyloid and tau accumulation.

**Conclusion:** This study provides evidence that the beneficial effects of LS7 on cognition are primarily through cerebrovascular pathways rather than direct influences on AD pathology.

## Background

While Alzheimer’s disease (AD) is the most common form of dementia, accounting for 60-70% of cases, it has been estimated that up to half of AD cases include a vascular component, culminating in mixed pathology.^1^ Based on neuropathology alone, approximately 26% of dementia cases have vascular dementia, but no AD pathology.^1^

Amyloid plaques and tau tangles are markers of AD pathology that can be observed decades before a clinical diagnosis. Similarly, markers of cerebrovascular disease (CBVD) appear long before the clinical diagnosis of vascular dementia and have been linked to cognitive decline.^2^ These markers include the volume of white matter hyperintensities (WMH) and reduced blood flow as measured by brain magnetic resonance imaging (MRI).

Poor cardiovascular health has been associated with cognitive decline, dementia, and WMH in late-life.^3,4^ Cardiovascular disease risk can be estimated according to the American Heart Association’s “Life Simple 7 ideal cardiovascular health recommendations” (LS7), which include smoking status, dietary intake, physical activity, body mass index (BMI), fasting glucose, cholesterol, and blood pressure.^5^

While cardiovascular disease risk has been associated with WMH and cognitive decline, its role in AD pathology is unknown.^6^ Here, we aimed to test the association between the LS7 score and markers of CBVD, dementia, and AD. A secondary aim was to determine the relevant time frame between the LS7 exposure and the outcomes. We know from the Whitehall 2 study that the impact of lifestyle factors on dementia can have a significant lag, with dementia incidence observed up to 25 years after the measured lifestyle factors.^4^ To account for this, we tested associations at different time periods (2-4 years, 4-6 years, and 6-8 years) between predictor and outcome. Additionally, we sought to determine if the LS7 score is associated with cognition through one or both pathways (CBVD and/or AD).

## Methods

### Study population

The Wisconsin Registry for Alzheimer’s Prevention (WRAP) cohort is enriched for individuals who have a parental history of AD and may be in the preclinical stages of the disease. The study consists of a longitudinal observational cohort started in 2001, with the initial follow-up beginning in 2005 and further visits performed on a biannual basis.^7^ This study was approved by the Institutional Review Board (IRB) at the University of Wisconsin and conducted in accordance with the principles of the Helsinki Declaration. Written informed consent was obtained from all participants. Approximately 80% of the participants who were enrolled at baseline have remained active in the study.^8^ In addition, continued recruitment and enrollment has occurred over the life of the study to increase the representation of minorities, including African Americans, Native Americans, Asians, and Hispanics. Participants were English speaking, age 40-65, and free of dementia at baseline. All participants in the study have undergone neuropsychological testing, clinical evaluation, behavioral risk factor assessment, and blood collection at each visit. A subset of participants underwent brain imaging to monitor for signs of AD and CBVD as of January 2024 and December 2023 respectively.

### Cerebrovascular disease imaging

A subset of participants completed at least one MRI, which provided measurements of WMH, white matter, grey matter, cerebrospinal fluid (CSF) volume, and arterial spin labeling (ASL) perfusion. WMH volume was measured via MRI, using 3D T2-weighted FLAIR scans and calculated from the total volume of WMH (cubic cm).^9^ To ensure equivalent signal sensitivity, only imaging collected using the following head coils was included in analysis: 48HAP, RM:Nova32channel, 8US TORSOPA, and 30AA+60PA. Percent WMH was calculated by dividing the total WMH volume by the total white matter volume, to account for varying white matter volume among participants, then multiplying by 100. Additionally, cerebral blood flow (CBF) was measured via ASL perfusion in both gray and white matter.

For the CBVD imaging outcomes, which included %WMH and ASL perfusion, normality of the residuals was assessed using QQ plots, histograms, and the Shapiro-Wilk test. The %WMH data showed skewness, and a cubic root transformation was applied to improve normality and stabilize variance, allowing the residuals to meet the assumptions of the regression model. In contrast, the ASL perfusion data were already normally distributed, so no transformations were applied, and the original data were used in the analysis.

### Alzheimer’s disease imaging

A subset of the participants completed at least one positron emission tomography (PET) paired with a T1-weighted MRI for anatomical reference. Multiple tracers were used during imaging to measure AD pathology. In this study, we were interested in the Pittsburgh compound B (PiB) tracer which assesses accumulation of beta-amyloid, and MK6240 which assesses accumulation of tau. The PiB index used in this study is an average of distribution volume ratios of cerebral amyloid within eight selected cortical regions of interest: angular gyrus, anterior cingulate gyrus, posterior cingulate gyrus, frontal medial orbital gyrus, precuneus, supramarginal gyrus, middle temporal gyrus, and superior temporal gyrus, as detailed in Sprecher, et al 2015.^10–12^ For tau, the Mayo Temporal Composite (MTC) used in this study is a bilateral volume-weighted composite of the parahippocampal gyrus, amygdala, temporal fusiform gyrus, inferior temporal gyrus, and the middle temporal gyrus. ^9^

The distributions of PiB index and MK6240 measurements were right-skewed, which is expected given the inclusion of both amyloid and tau negative vs positive individuals within the study sample. Transformations did not substantially improve normality. Consequently, they were analyzed in their original continuous form. Non-normal residuals were acknowledged as a limitation of the analysis, and this limitation is considered in the interpretation of results.

### Cognitive assessment

Cognitive composite scores (CCS), including Immediate Learning, Delayed Recall, Executive Function, and the Preclinical Alzheimer Cognitive Composite (PACC3), are widely used measures of cognitive function. Immediate Learning tests short-term memory, Delayed Recall tests long-term memory, Executive Function is a measure of cognitive processing, and PACC3 is indicative of global cognition.^13^ For the Executive Function and PACC3 CCS, an alternative composite score was created by the WRAP study team used for all visits in the analyses because the Stroop test (Executive Function) and Trail-Making and Digit Symbol tests (PACC3) were no longer possible during remote visits that occurred due to the COVID-19 pandemic. Instead, the Letter Fluency (‘C, F, L’) test was utilized as a substitute for those tests. All composite scores were transformed into z-scores by the WRAP study team, a commonly used technique in the field as detailed in Clark et al 2016.^13^

Cognitive composite scores were already standardized as z-scores with a mean of 0 and a standard deviation of 1. Visual inspection of residuals confirmed that, while slight deviations from normality were noted, the original z-score data maintained interpretability and were retained without further transformations.

### Life Simple Seven

Height was measured in centimeters and weight was measured in kilograms; BMI was calculated as weight in kilograms divided by height in meters squared. Participants were asked to relax in a quiet room prior to blood pressure (BP) readings; their BP was taken three times per each arm and then averaged to ensure accurate readings. Blood assays included fasting plasma glucose and serum total cholesterol.

Self-reported lifestyle factors included physical activity, smoking status, and diet. Participants were asked about the following in a typical week: intensity of exercise (mild, moderate, and hard), duration of the exercise in minutes, and frequency of the exercise per week. Current smoking habits were assessed by answering “Have you ever smoked cigarettes?” and “If, yes, in the past month, have you smoked any cigarettes at all?” Participants completed a set of 15 questions about diet, corresponding to each of the Mediterranean-Dash Intervention for Neurological Delay (MIND) diet categories (**Supplemental Table 1**).^14^

A total of 1775 participants were included in the WRAP November 2023 data freeze. 1231 participants had complete data for all LS7 components at the same visit. If there were multiple visits with complete LS7 data, the earliest visit was used.

*Scoring LS7.* Each of the seven LS7 components was assessed independently to provide a score of 0 (poor), 1 (intermediate), or 2 (optimal; **Table 1**). The LS7 score had a minimum of 0 and maximum of 14. The LS7 scoring criterion for diet was adjusted to reflect the data available in the WRAP cohort. In Sabia et al 2019,^4^ authors assessed diet based on fruit and vegetable consumption, plus high fiber bread. While the WRAP cohort contains data on fruit and vegetable consumption, high fiber bread was not included in the questionnaire and whole grains were used instead. Smoking history was determined by utilizing the longitudinal data of the WRAP study.

**Table 1.** Life Simple 7 definition of cardiovascular health metrics.

|  | Smoking | Diet* | Physical Activity Intensity (min/week) | BMI | Fasting Glucose | Blood Cholesterol | Systolic & Diastolic Blood Pressure |
| --- | --- | --- | --- | --- | --- | --- | --- |
| **Optimal (Score = 2)** | Never smoked<br>**OR**<br>Stopped >5 years ago | ≥2 servings of fruits and ≥2 of vegetables,<br>**PLUS**<br>≥1 serving of whole grains | ≥150 moderate<br>**OR**<br>≥75 hard<br>**OR**<br>≥150 moderate and hard | <25 | <100 mg/dL<br>Untreated | <200 mg/dL<br>Untreated | SBP <120 mm Hg and DBP <80 mm Hg<br>Untreated |
| **Intermediate (Score = 1)** | Stopped in past 5 years | ≥2 servings of fruits and ≥2 of vegetables,<br>**OR** ≥1 serving of whole grains | 1-149 moderate<br>**OR**<br>1-74 hard<br>**OR**<br>1-149 moderate and hard | 25 – 29.9 | <100 mg/dL Treated<br>**OR**<br>100-125 mg/dL | <200 mg/dL Treated **OR** 200-239 mg/dL | SBP <120 mm Hg and DBP <80 mm Hg Treated<br>**OR**<br>SBP 120-139 **OR** DBP 80-89 mm Hg |
| **Poor (Score = 0)** | Current Smoker | <2 servings of fruits and <2 of vegetables,<br>**PLUS** no whole grains | No moderate or hard | ≥30 | ≥126 mg/dL | ≥240 mg/dL | SBP ≥140 mm Hg<br>**OR** DBP ≥90 mm Hg |

\*Scoring criteria for each participant were according to the original LS7 except diet was assessed using Fruit, Vegetable, and Whole Grain consumption. Adapted from: Sabia et al 2019.<sup>4</sup>

At the biannual visits, each participant provides an update on their smoking status. We created a longitudinal smoking history measure that looked back at the 3 previous visits to identify whether they were a never smoker, previous smoker who stopped less than or greater than 5 years ago, or current smoker. All other criteria were consistent with the LS7 scoring in Sabia et al.^4^

### Outcome timing

This study used longitudinal data from the WRAP cohort, with outcomes collected over time as new imaging and diagnostic methods became available. Cognitive testing began in 2005, PET imaging for AD biomarkers was available from 2009, and MRI imaging for CBVD using the aforementioned head coils started in 2019. Each outcome—CCS, AD biomarkers, and CBVD measures—was analyzed separately.

Predictor data (for all LS7 components) were collected between 2014 and 2023. To ensure temporal separation, outcomes were only included if they occurred at least two years after the predictor data. Time intervals between predictor and outcome were categorized into 2-4 years, 4-6 years, and 6-8 years. Each time period was treated as a separate dataset, with the latest available outcome within each time window used for the analysis. The "Years to Outcome" variable was calculated as the difference between the participant’s age at the outcome visit and their age at the predictor visit. Participant counts for each dataset (time period) are shown in **Figure 1**. A minimum of 60 participants was required for analysis, and all outcomes met this criterion, except for the 8-10 year interval, which was subsequently excluded from the analyses. On average, there was just over one imaging outcome per individual, whereas the cognitive data were assessed at each visit.

**Figure 1:**
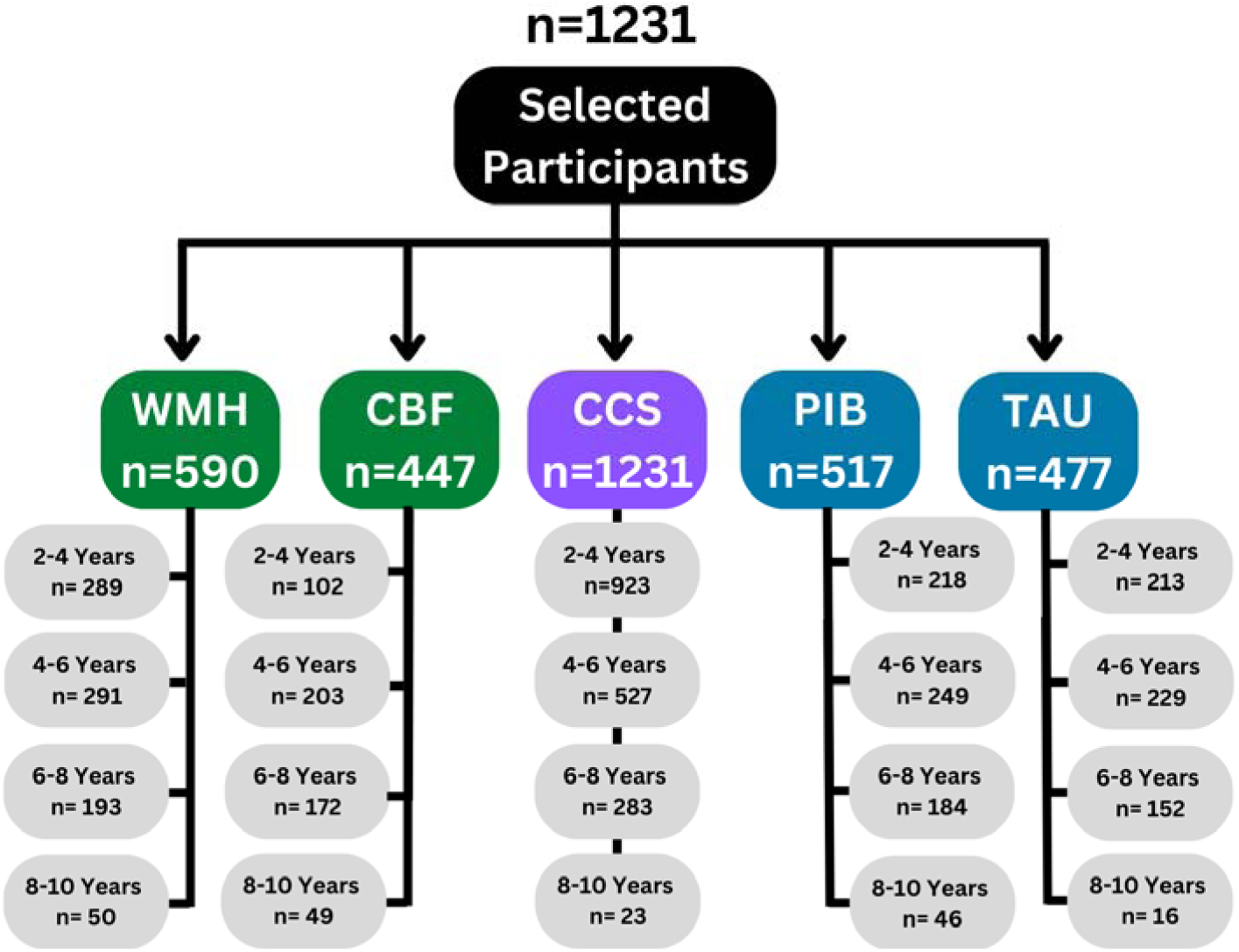
Sample sizes for the outcomes and time period between the predictor and outcome.

A total of 1231 WRAP participants had data available for all seven of the Life Simple Seven (LS7) components for at least one time period. The number of participants with each outcome is shown, including white matter hyperintensities (WMH), cerebral blood flow (CBF), cognitive composite scores (CCS), amyloid imaging (PIB), and tau imaging (TAU). Time periods were categorized based on the time between the predictor (LS7) and the outcome: 2-4 years, 4-6 years, 6-8 years, and 8-10 years. Each outcome and time period were analyzed as a unique dataset, with participant counts varying due to staggered enrollment, varying availability of outcome measures, and the structure of follow-up visits. Only the most recent observation for each participant within a given time period was included. CCS was the only outcome measured in all participants, whereas imaging outcomes (WMH, CBF, PIB, and TAU) were measured in a subset of the participants.

### Regression analysis

Generalized linear regression models were constructed using R studio package lme4.^15^ Separate models were run for each outcome: %WMH, ASL perfusion, Immediate Learning, Delayed Recall, Executive Function, PACC3, amyloid via PiB index, and tau via MK6240. All models controlled for sex, with males as the reference, and age, using age centered at 65 years. Models for cognitive outcomes additionally controlled for *APOE* ε4 count, practice effects (the number of times the participant completed cognitive testing), and education in years. The *APOE* ε*4* allele increases the risk of developing late onset AD, so it was included in the CCS and AD outcomes, however its contribution to CBVD is not well established and therefore it was not included in the WMH or ASL perfusion analyses.^16^ The predictor variable of interest for all models was the LS7 score. The significance threshold was predetermined to be set at p-value <0.05.

A key assumption of generalized linear models is that the residuals are approximately normally distributed to ensure the accuracy of parameter estimates and hypothesis testing. To assess this assumption, we conducted visual inspections of residuals using QQ plots and histograms, and performed statistical tests such as the Shapiro-Wilk test). When significant deviations from normality were detected, transformations (cubic root and log) were considered to stabilize variance and improve normality. Decisions regarding specific transformations and the handling of non-normal residuals are detailed in the outcome-specific sections above.

### Exploratory post-hoc mediation analysis

To explore the potential pathway through which LS7 affects cognitive outcomes, we conducted post-hoc mediation analyses. Mediation models included LS7 as the predictor, %WMH or ASL perfusion as mediators, and cognition (PACC3 and Delayed Recall) as outcomes. The timing of these variables was carefully aligned to ensure a logical progression: as described previously, the earliest visit with complete LS7 data was used, imaging mediators were measured 2-4 years later, and cognitive outcomes were assessed either concurrently with the imaging or up to two years afterward (ranging from 6 months before to 2 years after imaging). This requirement for the cognitive outcomes to be assessed at or after the imaging resulted in a slightly smaller sample size than was available for the 2-4 year time point for CBVD markers. Given the lack of association between LS7 and AD markers, mediation analyses with these markers (amyloid and tau) were conducted for completeness but were not expected to yield significant results. Indirect, direct, and total effects were estimated using the nonparametric bootstrap method with 1,000 simulations.

## Results

For all outcomes, we compared the magnitude of the beta coefficients for a one-point increase in the LS7 score to a one-year increase in age. This comparison was used to contextualize the impact of lifestyle as measured by the LS7 score against the well-established risk factor of age. This approach provides a clearer interpretation of how lifestyle improvements relate to age-associated risks.

### Participant characteristics

The participant characteristics are based on CCS and vary slightly based on the outcome of interest where CBVD and AD imaging are subsets of the CCS group. The majority of the WRAP cohort participants were female (67-78%) and non-Hispanic white (91-95%; **Table 2**). The cohort is highly educated with most participants having some college education (13 or more years). Due to the nature of recruitment, the WRAP cohort has been enriched for a parental history of AD and 35-37% had at least one *APOE* ε4 allele. The total LS7 score ranged from 3-14, with the mean and median being a score of 9 to 10 out of 14. This means that, in general, the cohort had a moderately healthy lifestyle. At the 2-4 year time period between predictor and outcome, the average age of participants at the outcome measurement was 67.1 for WMH, 65.8 for ASL perfusion, 66.6 for cognitive testing, 67.4 for PiB amyloid index, and 67.2 for MK6240 tau imaging.

**Table 2.**
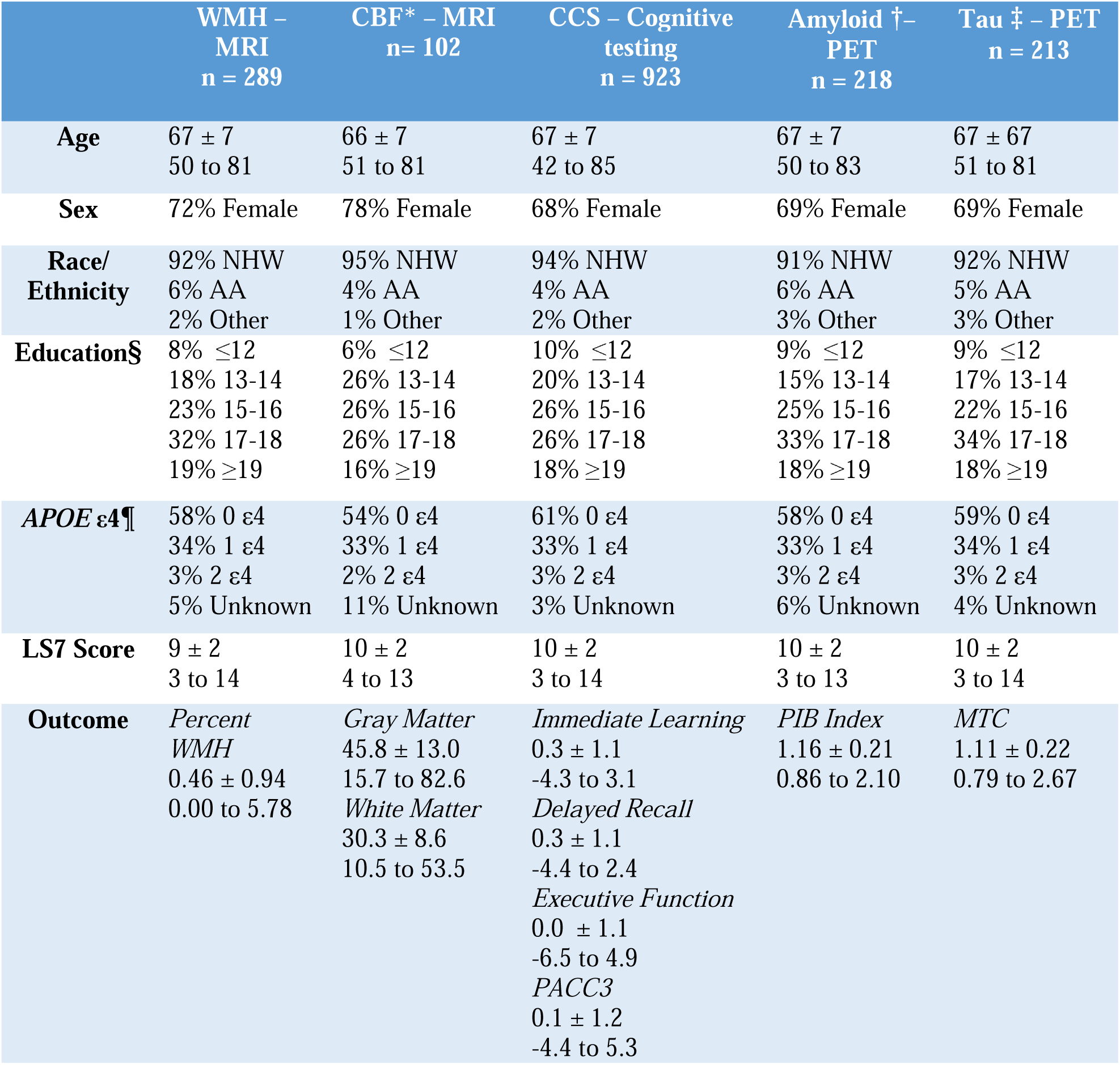
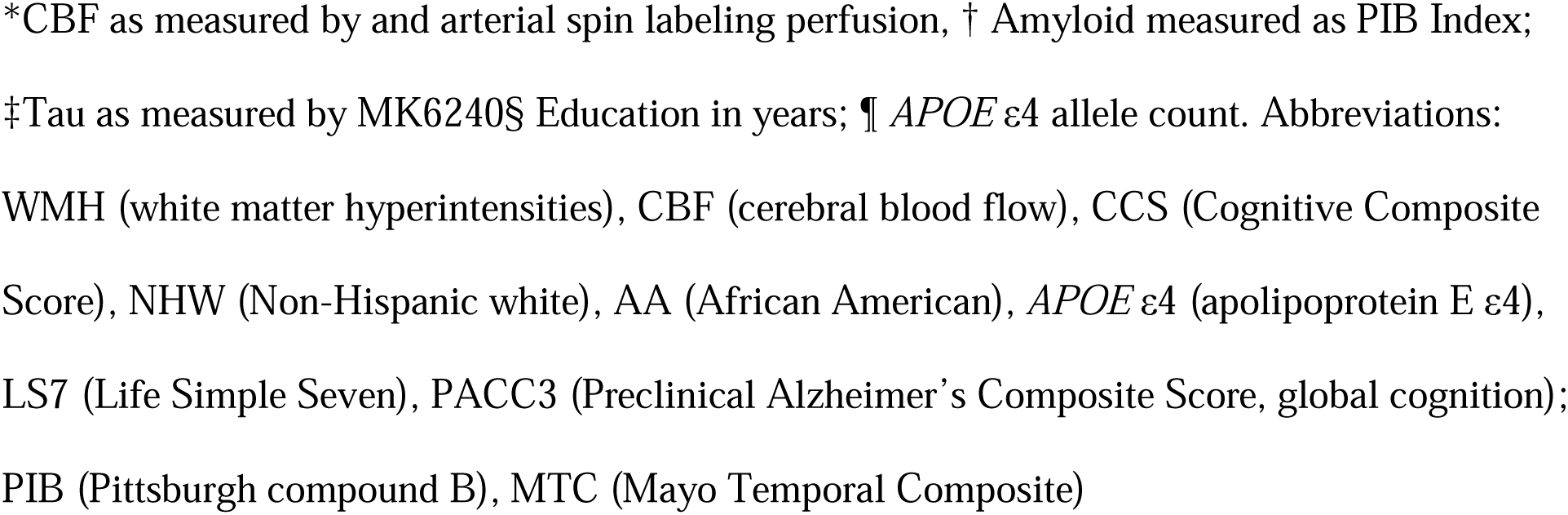
Characteristics of the participants in each biomarker dataset at 2-4 year time period.

|  | WMH –<br>MRI<br>n = 289 | CBF* – MRI<br>n= 102 | CCS – Cognitive<br>testing<br>n = 923 | Amyloid †–<br>PET<br>n = 218 | Tau ‡ – PET<br>n = 213 |
| --- | --- | --- | --- | --- | --- |
| **Age** | 67 ± 7<br>50 to 81 | 66 ± 7<br>51 to 81 | 67 ± 7<br>42 to 85 | 67 ± 7<br>50 to 83 | 67 ± 67<br>51 to 81 |
| **Sex** | 72% Female | 78% Female | 68% Female | 69% Female | 69% Female |
| **Race/**<br>**Ethnicity** | 92% NHW<br>6% AA<br>2% Other | 95% NHW<br>4% AA<br>1% Other | 94% NHW<br>4% AA<br>2% Other | 91% NHW<br>6% AA<br>3% Other | 92% NHW<br>5% AA<br>3% Other |
| **Education§** | 8% ≤12<br>18% 13-14<br>23% 15-16<br>32% 17-18<br>19% ≥19 | 6% ≤12<br>26% 13-14<br>26% 15-16<br>26% 17-18<br>16% ≥19 | 10% ≤12<br>20% 13-14<br>26% 15-16<br>26% 17-18<br>18% ≥19 | 9% ≤12<br>15% 13-14<br>25% 15-16<br>33% 17-18<br>18% ≥19 | 9% ≤12<br>17% 13-14<br>22% 15-16<br>34% 17-18<br>18% ≥19 |
| **APOE ε4¶** | 58% 0 ε4<br>34% 1 ε4<br>3% 2 ε4<br>5% Unknown | 54% 0 ε4<br>33% 1 ε4<br>2% 2 ε4<br>11% Unknown | 61% 0 ε4<br>33% 1 ε4<br>3% 2 ε4<br>3% Unknown | 58% 0 ε4<br>33% 1 ε4<br>3% 2 ε4<br>6% Unknown | 59% 0 ε4<br>34% 1 ε4<br>3% 2 ε4<br>4% Unknown |
| **LS7 Score** | 9 ± 2<br>3 to 14 | 10 ± 2<br>4 to 13 | 10 ± 2<br>3 to 14 | 10 ± 2<br>3 to 13 | 10 ± 2<br>3 to 14 |
| **Outcome** | *Percent*<br>*WMH*<br>0.46 ± 0.94<br>0.00 to 5.78 | *Gray Matter*<br>45.8 ± 13.0<br>15.7 to 82.6<br>*White Matter*<br>30.3 ± 8.6<br>10.5 to 53.5 | *Immediate Learning*<br>0.3 ± 1.1<br>-4.3 to 3.1<br>*Delayed Recall*<br>0.3 ± 1.1<br>-4.4 to 2.4<br>*Executive Function*<br>0.0 ± 1.1<br>-6.5 to 4.9<br>*PACC3*<br>0.1 ± 1.2<br>-4.4 to 5.3 | *PIB Index*<br>1.16 ± 0.21<br>0.86 to 2.10 | *MTC*<br>1.11 ± 0.22<br>0.79 to 2.67 |

\*CBF as measured by and arterial spin labeling perfusion, † Amyloid measured as PIB Index; ‡Tau as measured by MK6240§ Education in years; ¶ *APOE* ε4 allele count. Abbreviations: WMH (white matter hyperintensities), CBF (cerebral blood flow), CCS (Cognitive Composite Score), NHW (Non-Hispanic white), AA (African American), *APOE* ε4 (apolipoprotein E ε4), LS7 (Life Simple Seven), PACC3 (Preclinical Alzheimer's Composite Score, global cognition); PIB (Pittsburgh compound B), MTC (Mayo Temporal Composite)

### LS7 and CBVD

*Percent WMH.* At all time periods, an increase in age was associated with a higher percent of WMH (**Figure 2a**, **Supplemental Table 2**). The LS7 was significant at 2-4 and 6-8 years between predictor and outcome, with an effect size for one point of the LS7 score comparable to one year of age. As noted earlier, this comparison highlights how improvements in LS7 scores relate to the established risk of aging.

**Figure 2:**
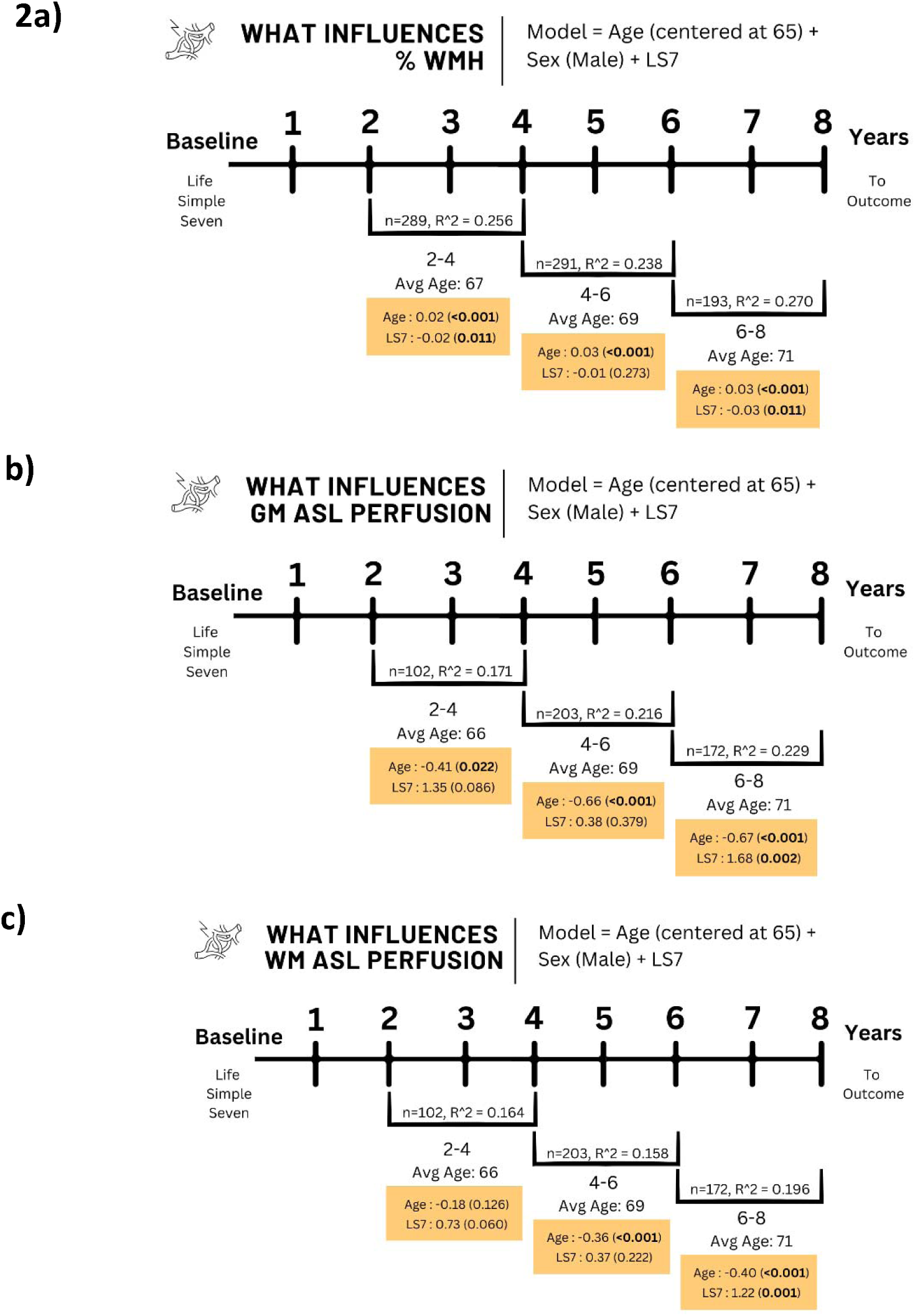
Cerebrovascular disease associations with Life Simple Seven (LS7) score.

*ASL Perfusion.* As expected, at all time-periods, an increase in age was associated with lower blood flow in both the white matter and gray matter (**Figure 2b-c, Supplemental Table 2**). A higher LS7 score was significantly associated with a higher perfusion level at 6-8 years, with an effect size from a one-unit improvement in the LS7 score similar to that of 2.5-3 years of age younger; the effect sizes in the 2-4 and 4-6 year time-periods were in the same direction, but not statistically significant.

Generalized linear model results for a) cubic root of percent white matter hyperintensities (%WHM), b-c) arterial spin labeling (ASL) perfusion measuring cerebral blood flow in ml/min/100 g for b) white matter and c) gray matter. All models controlled for age (centered at 65 years) and sex, with the LS7 score as the primary variable of interest. The sample size (n), variance (R^2^), and average age are provided for each time period (2-4 years, 4-6 years, and 6-8 years between predictor and outcome). Beta estimates (β) and p-values are presented for LS7 and age, which was included in the models to provide a benchmark for understanding the magnitude of LS7’s beta estimate; all other variables are shown in **Supplemental Table 2**. For %WMH, the cubic root transformation was applied to stabilize variance, with the raw calculation as follows %WMH = (WMH lesion volume / total white matter volume) x 100%.

### LS7 and Cognition

*PACC3 and Delayed Recall.* As expected, an increase in age was significantly associated with a decrease in z-scores for PACC3 and Delayed Recall at all three time points (**Figure 3a-b**, **Supplemental Table 3**). A higher LS7 score was significantly associated with higher PACC3 and Delayed Recall z-scores at 2-4 years, with an effect size from a one-unit improvement in the LS7 score nearly equivalent to that of one year of age younger; the effect sizes in the 4-6 and 6-8 year time-periods were in the same direction, but not statistically significant.

**Figure 3:**
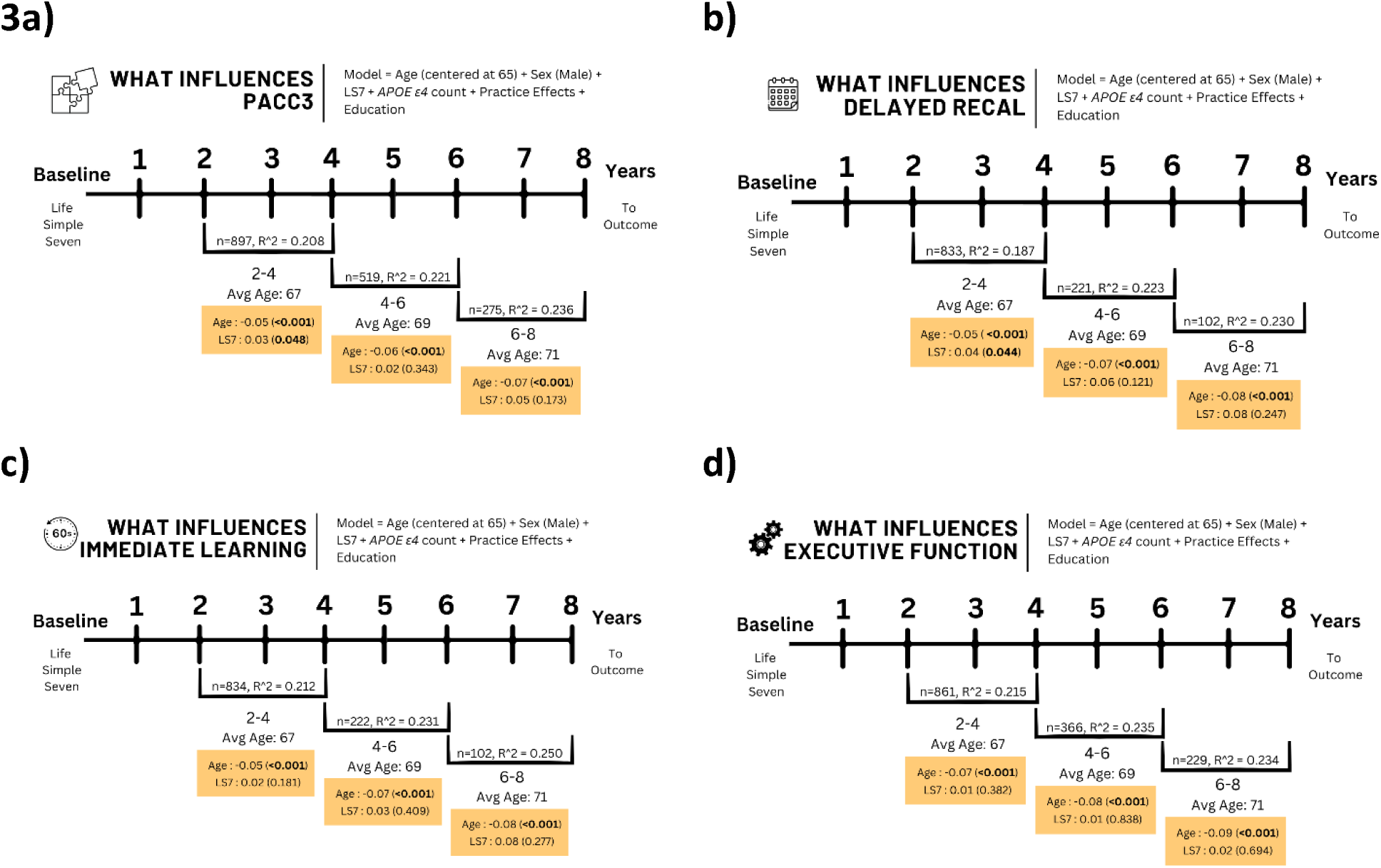
Cognitive composite score associations with Life Simple Seven (LS7) score.

#### Immediate Learning and Executive Function

An increase in age was significantly associated with a decrease in Immediate Learning and Executive Function z-scores at all three time points (**Figure 3c-d**, **Supplemental Table 3**). The LS7 was not significantly associated with these cognitive composite scores at any of the time points.

Generalized linear model results for z-scores of a) Pre-Clinical Alzheimer’s Cognitive Composite score 3 (PACC3), b) Delayed Recall, c) Immediate Learning, and d) Executive Function. All models controlled for age (centered at 65 years), sex, race/ethnicity, *APOE* ε*4* allele count, practice effects, education in years, with the LS7 score as the primary variable of interest. The sample size (n), variance (R^2^) and average age are provided for each time period (2-4 years, 4-6 years, and 6-8 years between predictor and outcome). Beta estimates (β) and p-values are presented for LS7 and age, which was included in the models to provide a benchmark for understanding the magnitude of LS7’s beta estimate; all other variables are shown in **Supplemental Table 3**.

### LS7 and AD markers

#### PiB index

The PiB index is a biomarker of amyloid accumulation as plaques in between the neurons; higher values indicate more amyloid accumulation and more AD pathology. Age and *APOE* ε4 allele count were the only significant contributors to the PiB index (**Figure 4a, Supplemental Table 4**).

**Figure 4:**
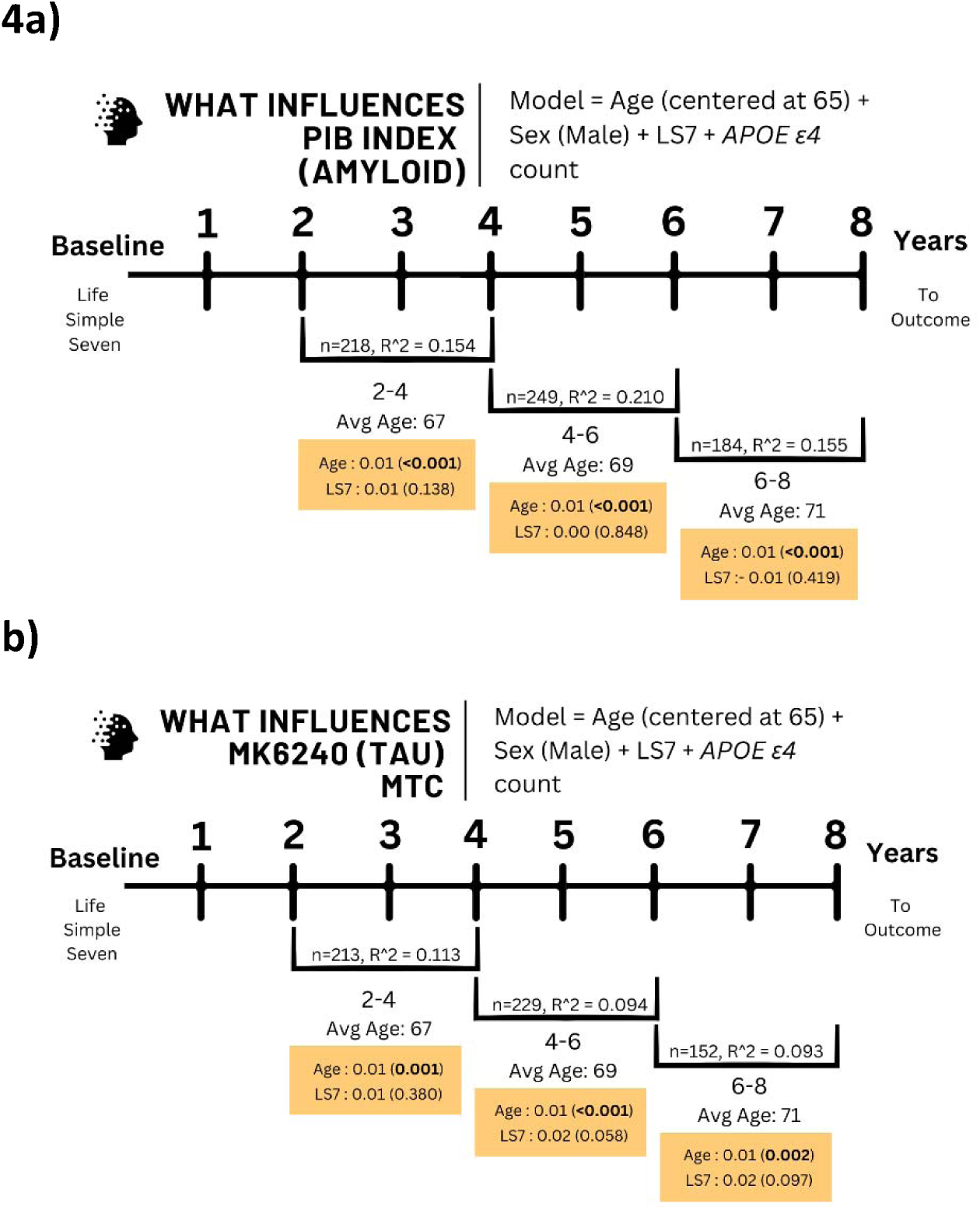
Alzheimer’s disease imaging associations with Life Simple Seven (LS7) score.

#### Tau MTC

The MTC is a biomarker of tau aggregation as tau tangles in four critical regions of the brain; higher values indicate more tau aggregation and more AD pathology. Age and *APOE* ε4 allele count were the only significant contributors to the MTC (**Figure 4b, Supplemental Table 4**).

Generalized linear model results for a) PIB index (amyloid) and b) Mayo Temporal Composite (MTC). All models controlled for age (centered at 65 years), sex, and *APOE* ε*4* allele count with the Life Simple Seven (LS7) score as the primary variable of interest. The sample size (n), variance (R^2^) and average age are provided for each time period (2-4 years, 4-6 years, and 6-8 years between predictor and outcome). Beta estimates (β) and p-values are presented for LS7 and age, which was included in the models to provide a benchmark for understanding the magnitude of LS7’s beta estimate; all other variables are shown in **Supplemental Table 4**.

### Mediation analysis

The results indicated that %WMH (cubic root, n=195) mediated 19.5% of the total effect of LS7 on PACC3, with the indirect and total effects being marginally significant (**Supplemental Figure 1a**). Similarly, for Delayed Recall, %WMH mediated 18.0% of the total effect of LS7, with the indirect and total effects being marginally significant (**Supplemental Figure 1b**). The mediation analysis for ASL perfusion (gray and white matter; n=57) did not show a significant indirect (mediation) effect on cognition, likely reflecting the limited sample size (**Supplemental Figure 1c-f**). For AD markers, PIB index and MK6240 MTC (n=148 and 147, respectively), no significant mediation was observed (**Supplemental Figure 2**). These findings support the hypothesis that LS7’s beneficial effects on cognition may be partly attributed to its impact on cerebrovascular health, emphasizing the importance of lifestyle modifications for dementia risk reduction.

## Discussion

In this longitudinal study involving 1231 participants from the WRAP cohort, we investigated the relationship between cardiovascular health, as measured by the LS7 score, and various indicators of brain health, including markers of CBVD and AD pathologies, and cognitive function. One of our primary goals was to determine the relevant time frame between the exposure (LS7 score) and outcomes. We found that higher LS7 scores, indicative of better cardiovascular health, were significantly associated with lower %WMH 2-4 and 6-8 years later, and higher CBF (ASL perfusion) 6-8 years later. Additionally, improved LS7 scores correlated with better cognitive performance in PACC3 and Delayed Recall composite scores 2-4 years later. These results highlight that the effect of cardiovascular health on brain health may be observed at different follow-up periods, with CBVD markers showing short-term and mid-term associations, while cognitive outcomes showed only short-term associations, although this finding requires replication in an independent sample. No significant associations were found between LS7 scores and AD markers, such as amyloid and tau accumulation, suggesting that the impact of LS7 on cognition is associated with changes in cerebrovascular health rather than direct influences on AD pathology. We also explored whether LS7 score was associated with cognition through one or both pathways (CBVD and AD). A post-hoc mediation analysis demonstrated that only the CBVD marker, %WMH, acted as a potential mediator between lifestyle and cognition.

Our findings align with previous research, reinforcing the role of cardiovascular health in maintaining cognitive function and reducing cerebrovascular disease risk. Wei et al. (2022) reported higher LS7 scores were associated with better scores on cognitive function tests in the NHANES (National Health and Nutrition Examination Survey) study,^17^ supporting our results of better cognitive outcomes with higher LS7 scores. Similarly, Liu et al. (2022), found that lower LS7 scores were associated with greater odds of cerebral small vessel disease in the PRECISE (Polyvascular Evaluation for Cognitive Impairment and Vascular Events) study.^18^ These results align with our findings that higher LS7 scores were associated with lower %WMH and better ASL perfusion, indicating healthier cerebrovascular function.

In regard to AD imaging, Gottesman et al. (2017) reported that midlife modifiable risk factors, such as smoking, diabetes, prehypertension, and hypertension, were associated with an increased risk of amyloid deposition and dementia, emphasizing the importance of addressing vascular health in midlife to mitigate long-term risks.^19^ While there are currently no publications focusing on the LS7 and AD imaging, Zhao et al. (2022) found that higher LS7 scores were associated with more favorable CSF AD biomarkers in the CABLE (Chinese Alzheimer’s Biomarker and LifestylE) study, particularly among *APOE* ε4 non-carriers, reporting higher (healthier) Aβ42/40 ratios and lower (healthier) pTau-181 associated with an increase in the LS7 score.^20^ This difference suggests that the relationship between LS7 and AD pathology may vary by biomarker type and participant characteristics, such as *APOE* status, which we did not stratify on due to limitations in sample size.

The LS7 includes seven easily accessible lifestyle metrics including diet, physical activity, smoking status, blood pressure, blood sugar, cholesterol, and BMI, which makes it ideal for implementation in a clinical setting. However, this is not the only lifestyle risk score in the current literature for dementia. Other risk scores have been developed with dementia in mind, including the CAIDE, LIBRA, ANU-ADRI, and CogDrisk scores.^21–25^ These scores share common elements, such as BMI, cholesterol, blood pressure, blood sugar, smoking, and physical activity, but differ in their inclusion of diet, cognitive/social activities, and additional diagnoses like depression, insomnia, and traumatic brain injury. Our study’s focus on LS7 highlights the importance of cardiovascular health in reducing dementia risk while offering a streamlined approach to intervention. Future research will consider the recently updated Life Essential Eight (LE8) from 2022, which incorporates sleep and refines the LS7 components.^26^

Age remains an established risk factor for CBVD, cognitive decline, and AD, with each year over age 65 contributing to increased risk.^27–29^ Consistent with this, our results showed age was significantly associated with poorer brain health across all outcomes. Notably, a one-unit increase in LS7 score was associated with an effect similar to being one year younger for %WMH at 2-4 and 6-8 years, and 2.5-3 years younger for CBF at 6-8 years. We did not see this association for %WMH at the 4-6 year time period, however this may be attributed to the wide CI overlapping with 0, and a lower R^2^ than the other two time periods. The differences between WMH and CBF, may be attributed to the nature of the two measures. WMH typically form due to ischemia, stroke, or microbleeds, meaning that they occur at a given point in time due to a specific event or as a result of chronically reduced blood flow, whereas CBF is more of a gradient where there can be a gradual reduction overtime, even among healthy adults. ^29–31^ Strengths of this study include a comprehensive set of outcomes with relatively large sample sizes. Few studies incorporate both CBVD and AD markers alongside cognition within the same cohort. Here we include both markers of vascular dementia and AD pathology along with cognition within the same cohort. A notable limitation was the small sample size at some time points, which impacted the significance of consistent effect sizes. Additionally, limited racial and ethnic diversity (6% non-Hispanic white) limited the generalizability of our findings. Future recruitment efforts aim to increase diversity in the WRAP cohort for more inclusive research.

Moreover, longitudinal data collection over time will address these limitations, increasing sample size and providing further follow-up visits to examine changes in LS7 scores and their long-term impact on brain outcomes.

In summary, our study demonstrates that better cardiovascular health, as measured by LS7 score, is associated with favorable CBVD and cognitive outcomes, but not with AD imaging markers. Higher LS7 scores correlated with a reduced risk of %WMH, improved CBF, and better cognitive performance, suggesting that cardiovascular health improvements are likely linked to reduced risk through the vascular pathway. This finding underscores the clinical significance of targeting cardiovascular health, as over 50% of dementia cases include a vascular component and could potentially benefit from diet and lifestyle modifications aimed at enhancing cardiovascular health.

Finally, given that multiple outside factors contribute to the risk of WMH, our ability to account for 25% of the variance underscores the clinical relevance of our findings and the potential for targeted intervention. Although AD pathology is multifaceted and influenced by a complex interplay of factors, our results highlight that the vascular contributions to dementia risk can be meaningfully addressed through lifestyle changes. Furthermore, while aging remains an unmodifiable risk factor, promoting strategies that reduce the formation of WMH and support CBF offers a practical and impactful approach for aging adults to mitigate the risk of dementia and cognitive decline.

## Author Contributions

Diandra N Denier-Fields (Conceptualization; Data Curation; Methodology; Formal analysis; Investigation; Writing - Original Draft; Writing - Review & Editing; Visualization), Ronald E Gangnon (Conceptualization; Methodology), Leonardo A Rivera-Rivera (Data Curation; Resources; Writing - Review & Editing), Tobey J Betthauser (Data Curation; Resources; Writing - Review & Editing), Barbara B Bendlin (Writing - Review & Editing), Sterling C Johnson (Funding acquisition), Corinne D Engelman (Conceptualization; Methodology; Writing - Review & Editing; Supervision; Project administration; Funding acquisition)

## Data Availability

The data underlying this article can be requested through the WRAP Application for Resources link on the following website: https://wrap.wisc.edu/data-requests-2/ (released on November 2023).

## Acknowledgements

The authors especially thank the WRAP participants and staff for their contributions to the studies. Without their efforts, this research would not be possible.

## Consent and Ethics

All participants provided signed informed consent before participation. All ethical guidelines were met, and IRB approvals have been obtained.

## Data Availability

The data supporting the findings of this study are available on request from the Wisconsin Registry for Alzheimer’s Prevention (https://wrap.wisc.edu/data-requests-2/). The data is not publicly available due to privacy restrictions.

