## Supplemental Tables 1-4 for "Evaluating Life Simple Seven’s influence on brain health outcomes: The intersection of lifestyle and dementia"

*Supplemental Table 1: Mediterranean-Dash Intervention for Neurological Delay (MIND) dietary questionnaire*

| Question | Frequency |
| --- | --- |
| 1) How many tablespoons of olive oil do you consume **per day** (including that used in salad dressings and sauteing? | _____ T per day |
| 2) How many servings of green leafy vegetables do you eat **each day**, such as spinach, kale, greens, romaine? (1cup for leafy, ½ cup for cooked/raw chopped) | _____ per day |
| 3) How many servings (1/2 cup) of other types of vegetables do you eat **each day** (e.g. broccoli, carrots, peas, onions, green/red peppers, celery, string beans, tomatoes, yams, squash, eggplant)? | _____ per day |
| 4) How many servings of berries do you eat **each week** (e.g. strawberries, blueberries, raspberries)? | _____per week |
| 5) How many servings of red meat (steak, ham, roast), hamburger, hot dogs or sausages do you consume **each week**? (3oz) | _____per week |
| 6) How many servings of fish (not fried and not including shellfish) do you consume **each week**? (3oz) | _____per week |
| 7) How many servings of chicken (not fried) do you consume **each week**? (3oz) | _____per week |
| 8) How many servings of whole fat or regular cheese or cream cheese do you eat **each week**? | _____per week |
| 9) How many servings of butter or cream (half & half) do you consume **each day**? (serving = 1T) | _____ T per day |
| 10) How many servings of beans (1/2 cup) do you consume **each week**? | _____per week |
| 11) How often do you eat whole grain breads, pasta, or cereals **each day**? (1 slice bread, ¾ cup pasta/cereal) | _____ per day |
| 12) How often do you consume sweets, candy bars, pastries, cookies or cakes **per week?** | _____per week |
| 13) How many servings of nuts do you eat **each week**? (handful or ¼ cup – 1/3 cup) | _____per week |
| 14) How many times **per week** do you consume food from a fast food restaurant such as McDonald’s, Burger King, Denny’s, Dominos, Popeyes, Kentucky Fried Chicken? | _____per week |
| 15) How many servings of alcohol (12 oz beer, a 5 oz glass of wine, or 1 shot (1.5 oz) of liquor) do you drink **each day?** | _____ per day |

*Supplemental Table 2: Cerebrovascular Disease Model Results*

| 2-4 years post LS7 | 4-6 years post LS7 | 6-8 years post LS7 |
| --- | --- | --- |
| 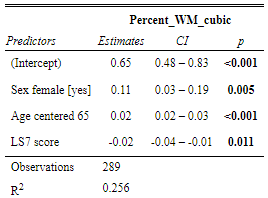 | 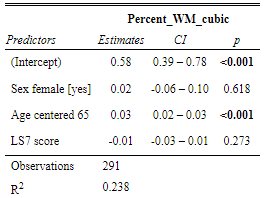 | 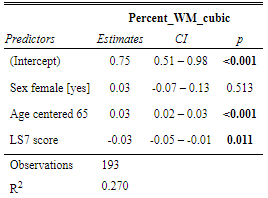 |
| 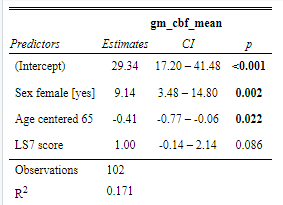 | 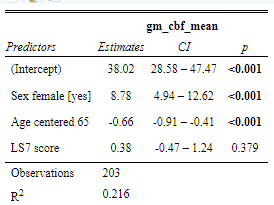 | 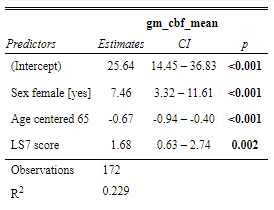 |
| 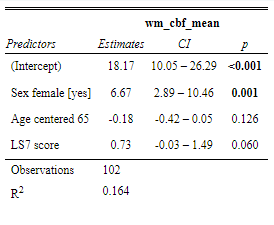 | 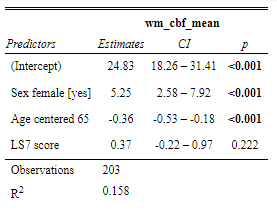 | 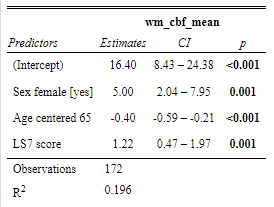 |

Abbreviations: LS7 (Life Simple Seven), percent_WM_cubic (the cubic root of percent white matter lesions), gm_cbf_mean (mean gray matter cerebral blood flow), wm_cbf_mean (mean white matter cerebral blood flow)

*Supplemental Table 3: Cognitive Composite Score Model Results*

| 2-4 years post LS7 | 4-6 years post LS7 | 6-8 years post LS7 |
| --- | --- | --- |
| 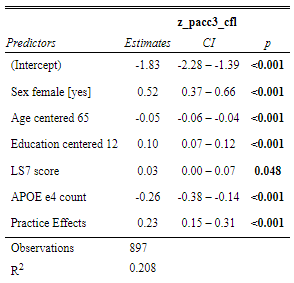 | 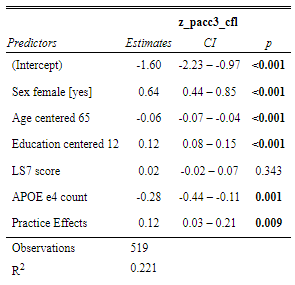 | 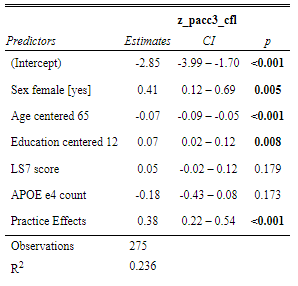 |
| 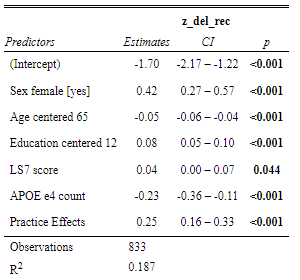 | 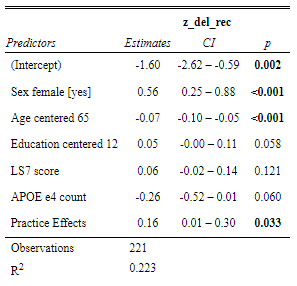 | 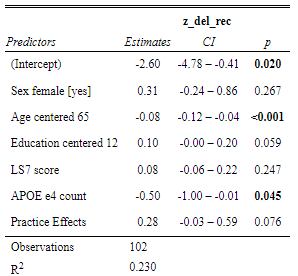 |
| 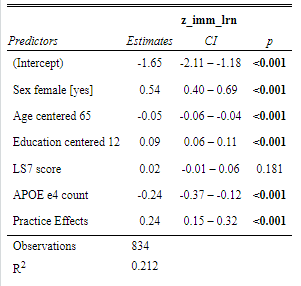 | 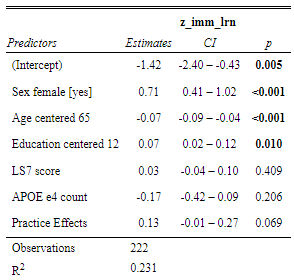 | 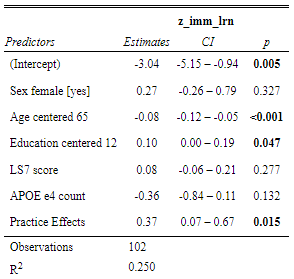 |
| 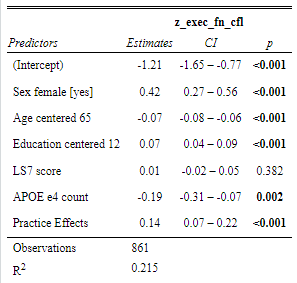 | 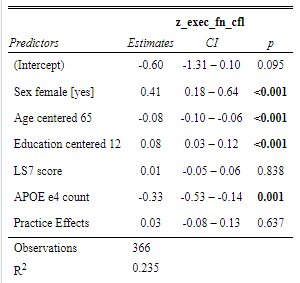 | 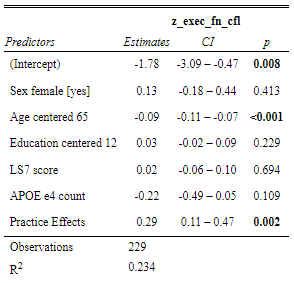 |

Abbreviations: LS7 (Life Simple Seven), z_pacc3_cfl (z-score for PACC3), z_del_rec (z-score for Delayed Recall), z_imm_lrn (z-score for Immediate Learning), z_exec_fn_cfl (z-score for Executive Function), education centered 12 (years of education based on high school graduate), practice effects (number of times participant has retaken the test)

*Supplemental Table 4: Alzheimer’s Disease Model Results*

| 2-4 years post LS7 | 4-6 years post LS7 | 6-8 years post LS7 |
| --- | --- | --- |
| 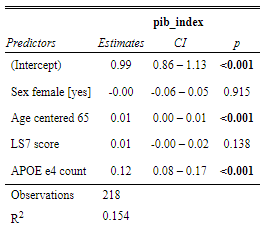 | 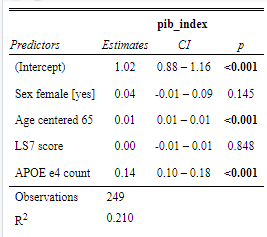 | 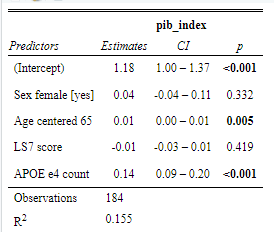 |
| 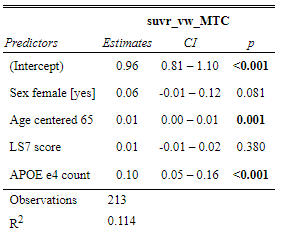 | 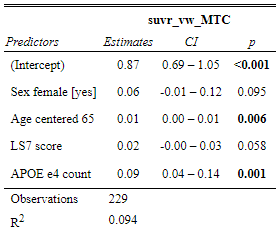 | 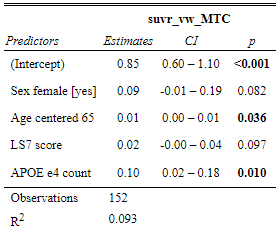 |

Abbreviations: LS7 (Life Simple Seven), pib_index (Pittsburgh compound B tracer for amyloid accumulation in brain), suvr_vw_MTC (MK6240 tracer for tau accumulation according to the Mayo Temporal Composite)
