## Supplemental Figures 1&2 for "Evaluating Life Simple Seven’s influence on brain health outcomes: The intersection of lifestyle and dementia"

| **1a)**  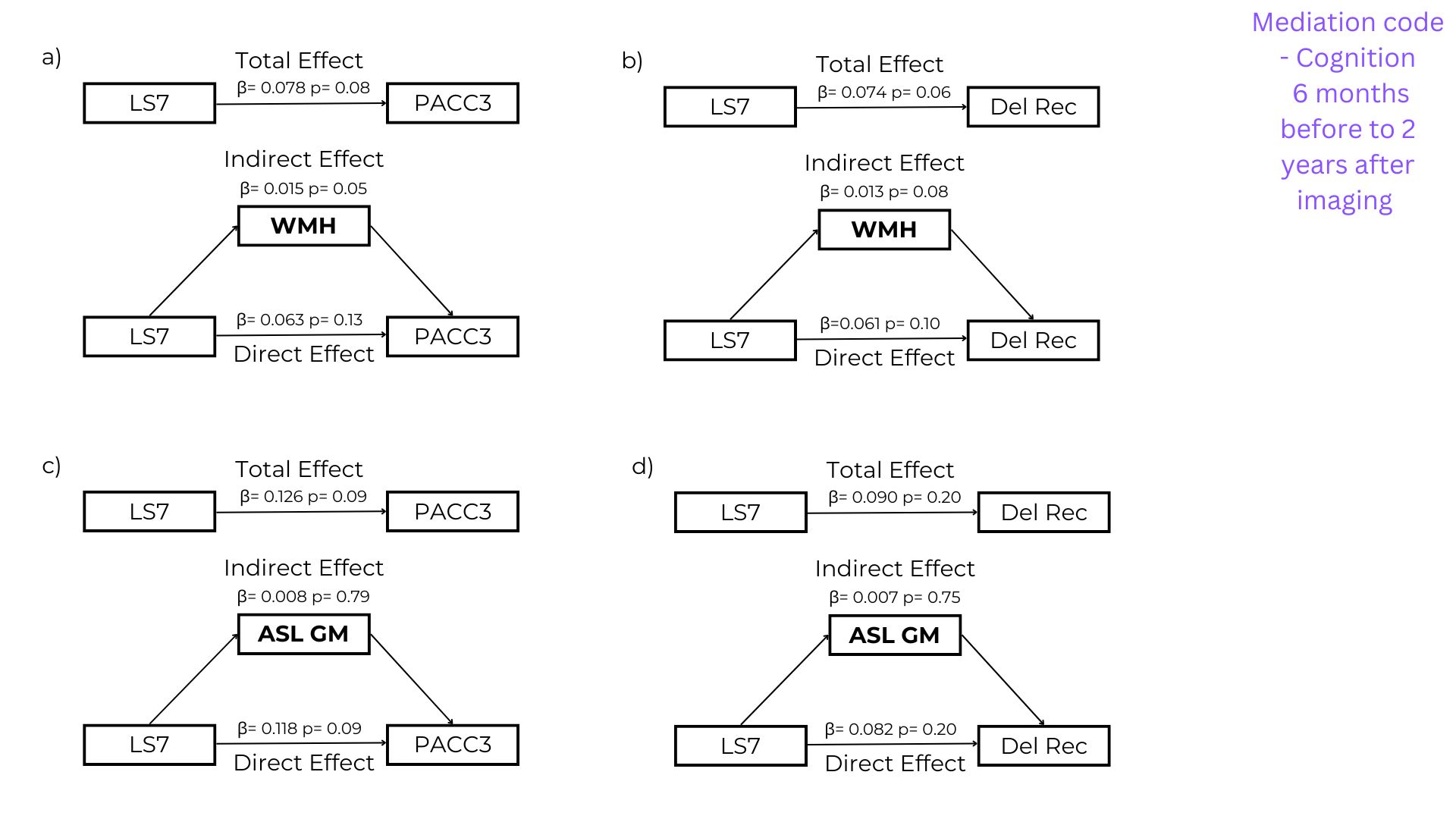 | **b)**  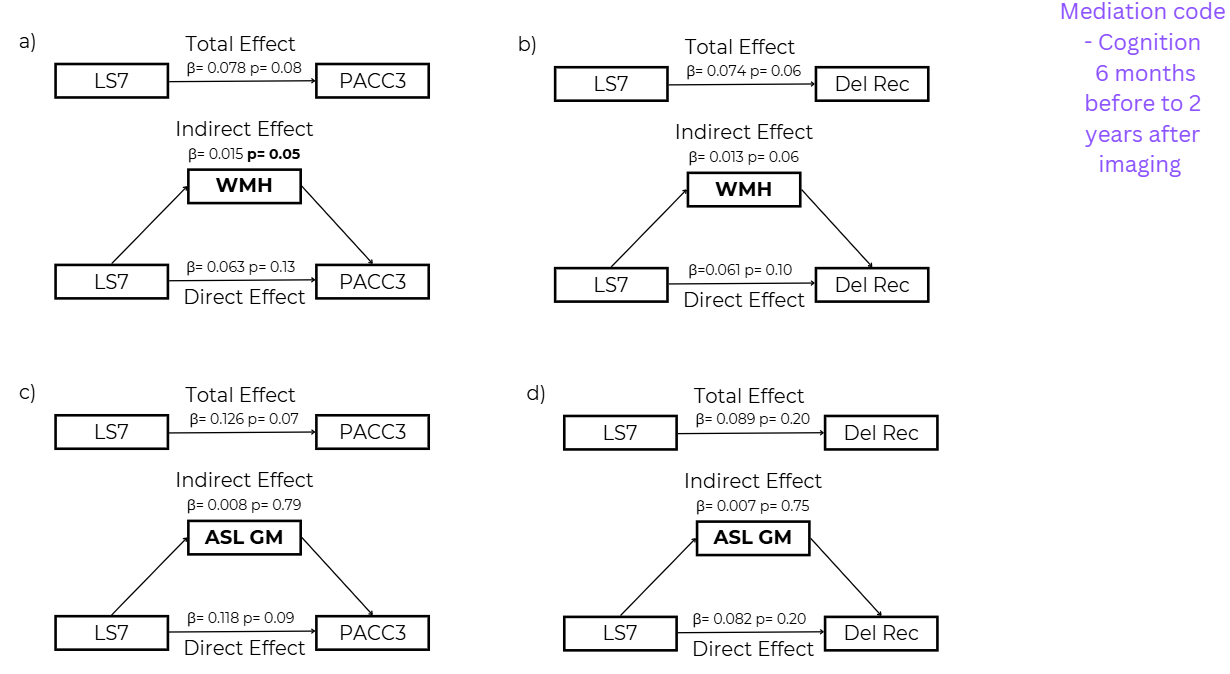 |
| --- | --- |
| **c)** | **d)** |
| 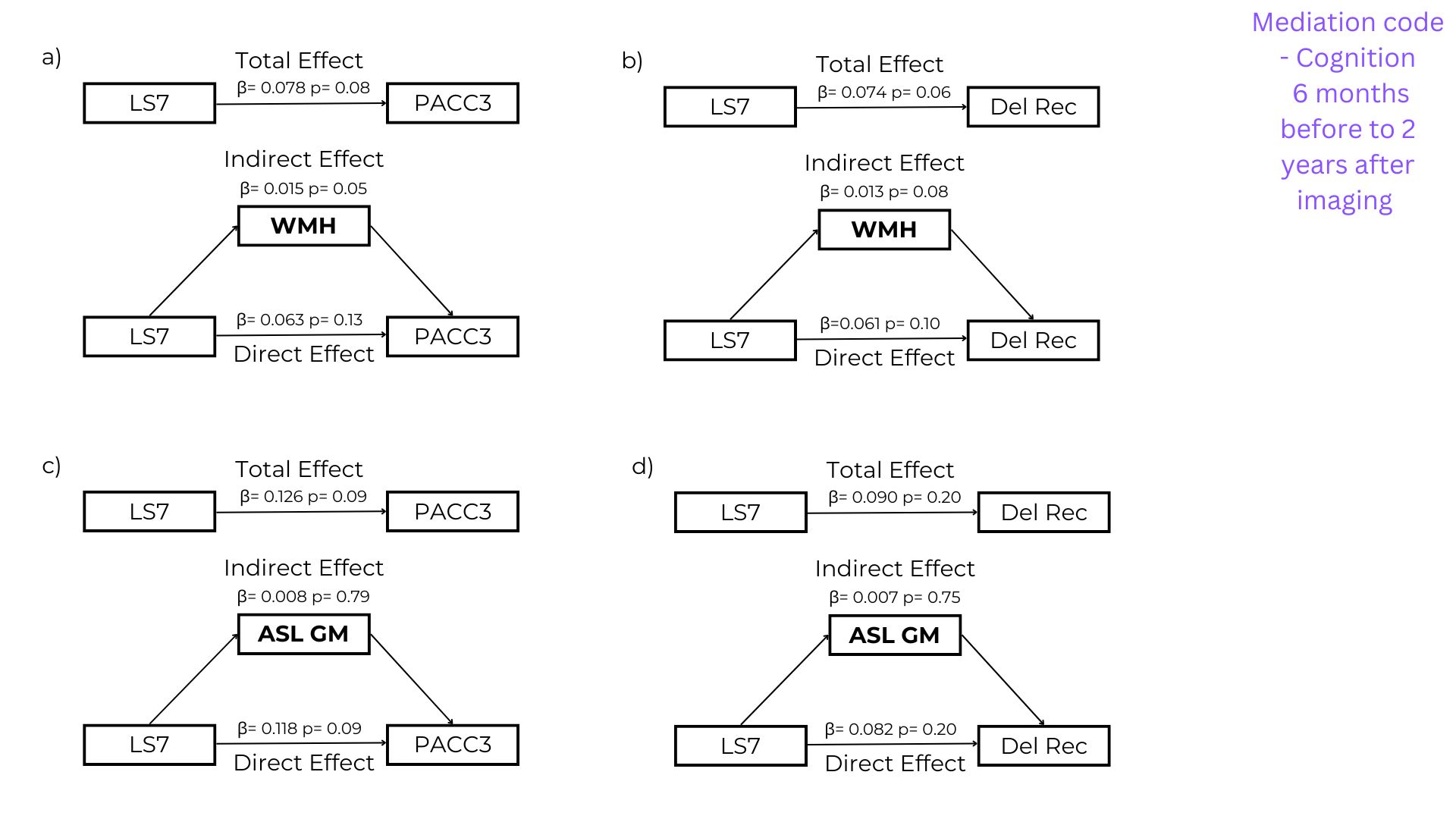 | 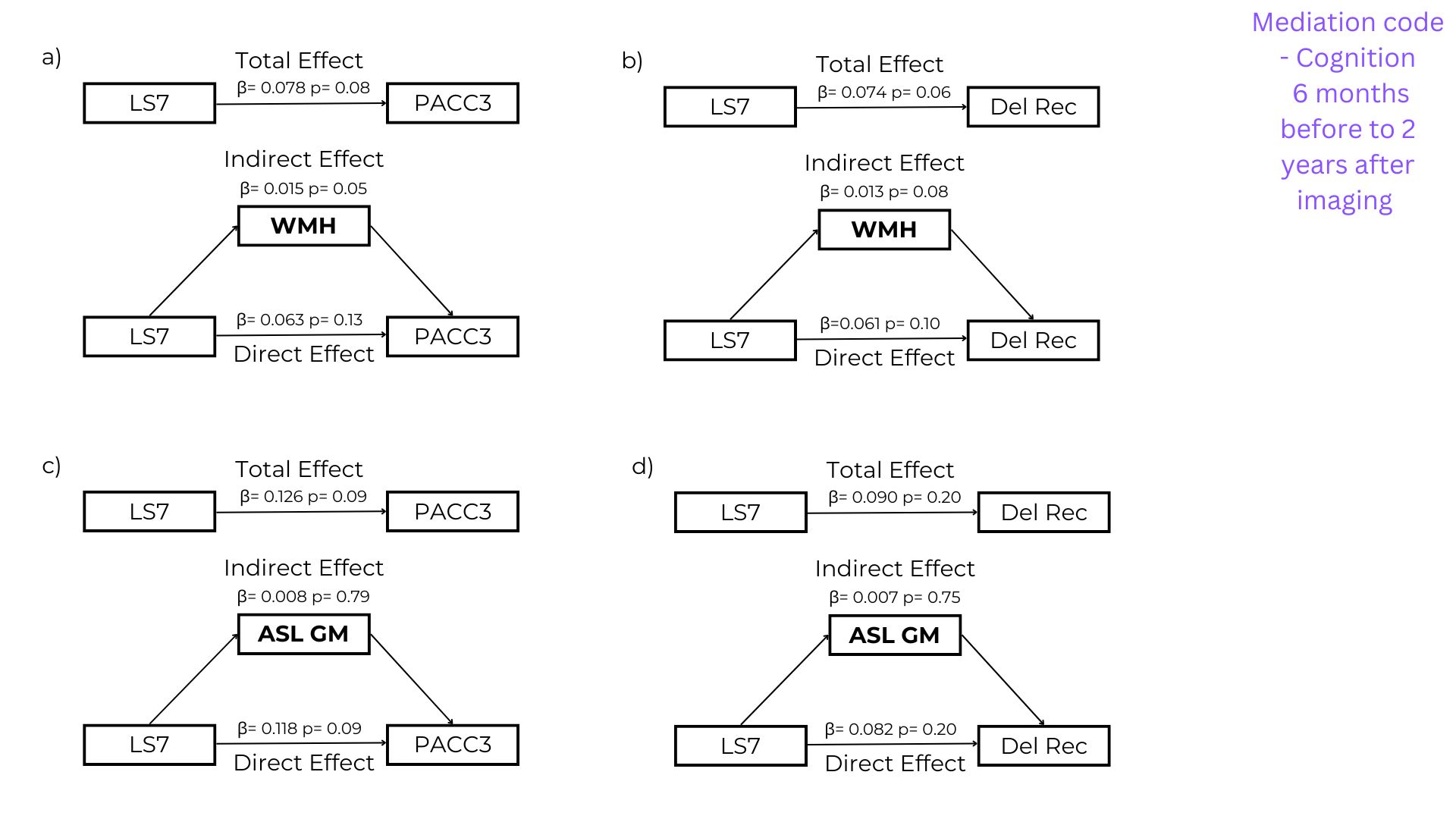 |
| **e)**  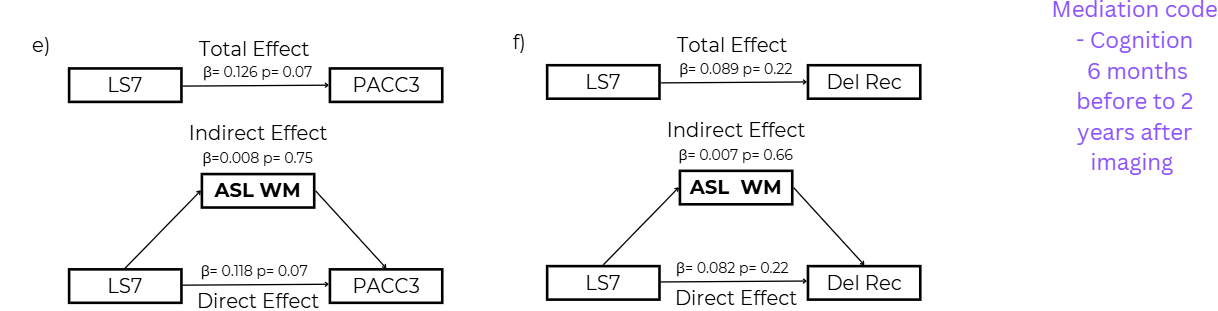 | **f)**  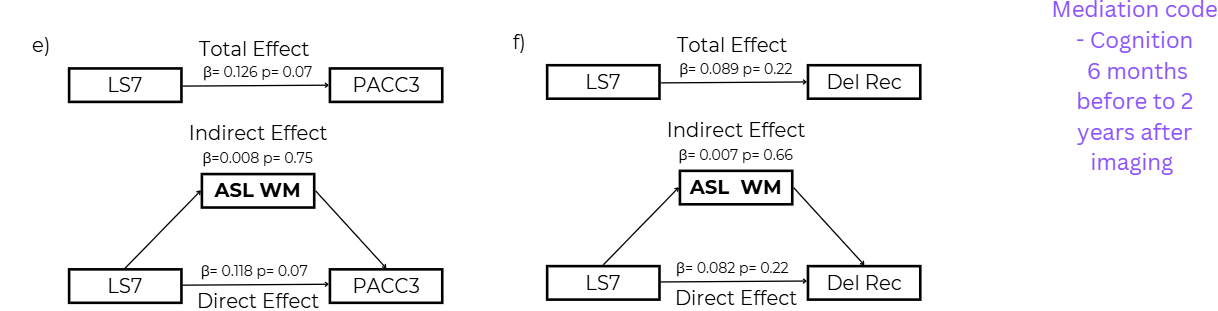 |

**Supplemental Figure 1: Mediation analysis of significant outcomes.** Mediation analysis results for the relationship between the Life Simple Seven (LS7) score, cerebrovascular and cognitive outcomes. The panels show the total effect, direct effect, and indirect effect (via the mediator) of LS7 on cognitive composite scores, Pre-Clinical Alzheimer’s Cognitive Composite score 3 (PACC3; a, c, and e) and delayed recall (Del Rec; b, d, and f). Mediators include cubic root-transformed %WMH (a, b; n=195) and ASL perfusion for gray matter (ASL GM; c, d; n=57) and white matter (ASL WM; e, f; n=57). Beta estimates (β) and p-values are provided for each pathway. Total effects reflect the overall association between LS7 and the cognitive composite score, indirect effects indicate the pathway through the mediator (cerebrovascular marker), and direct effects show the relationship between LS7 and the outcome independent of the mediator. The results underscore the complexity of the relationships between cardiovascular health, cerebrovascular pathways, and cognitive performance.

| **2a)**  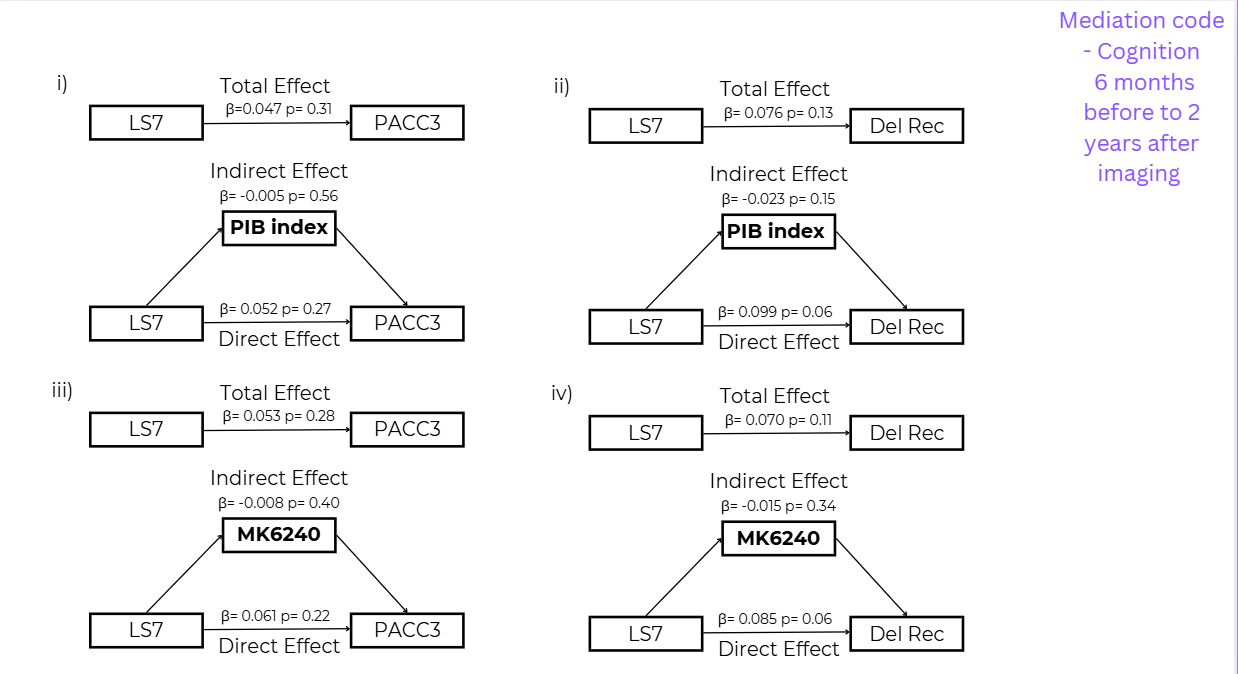 | **b)**  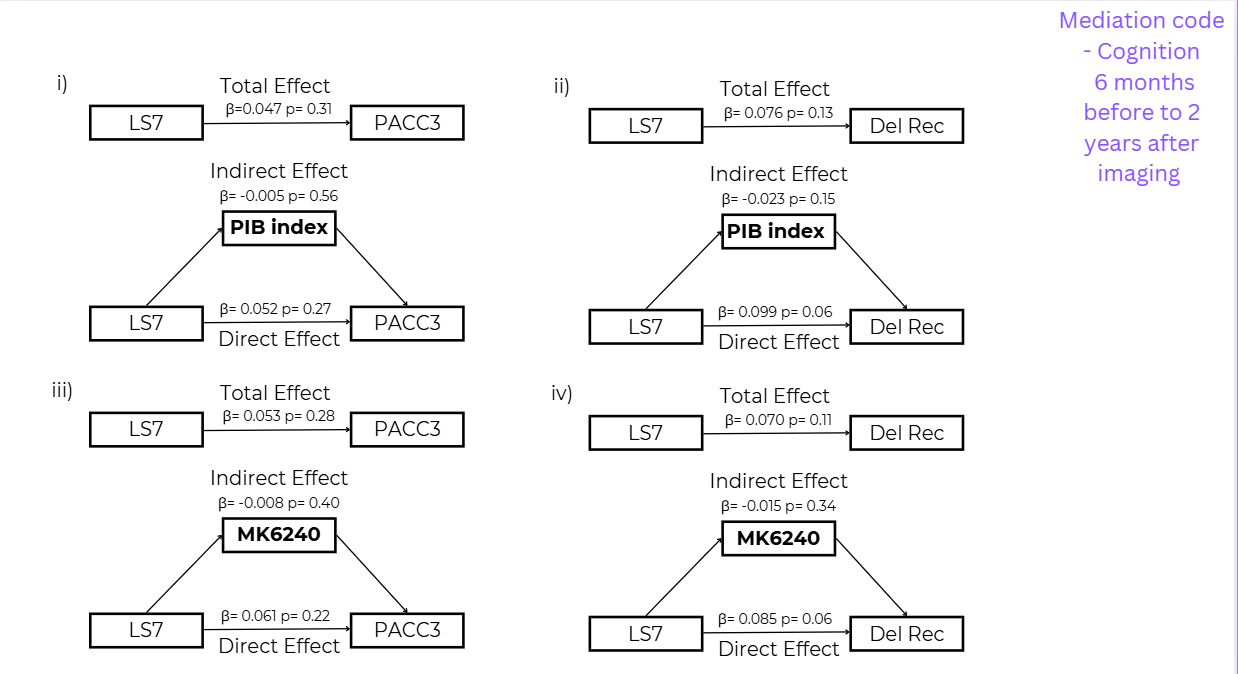 |
| --- | --- |
| **c)** | **d)** |
| 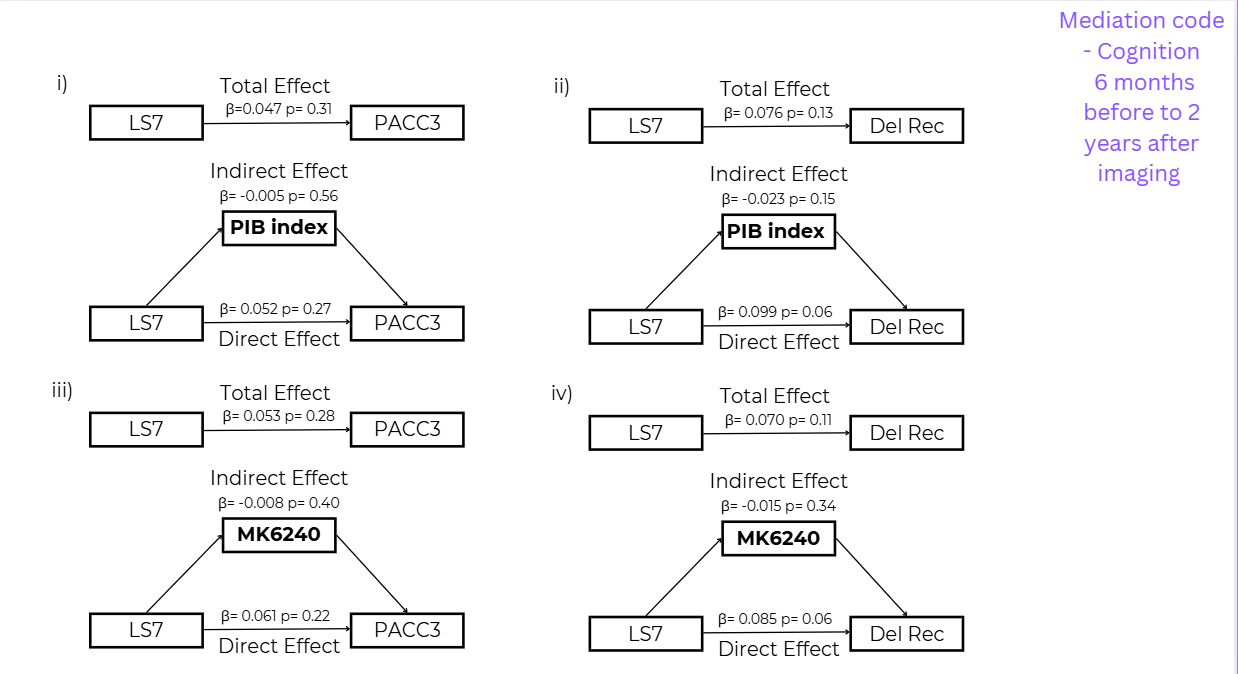 | 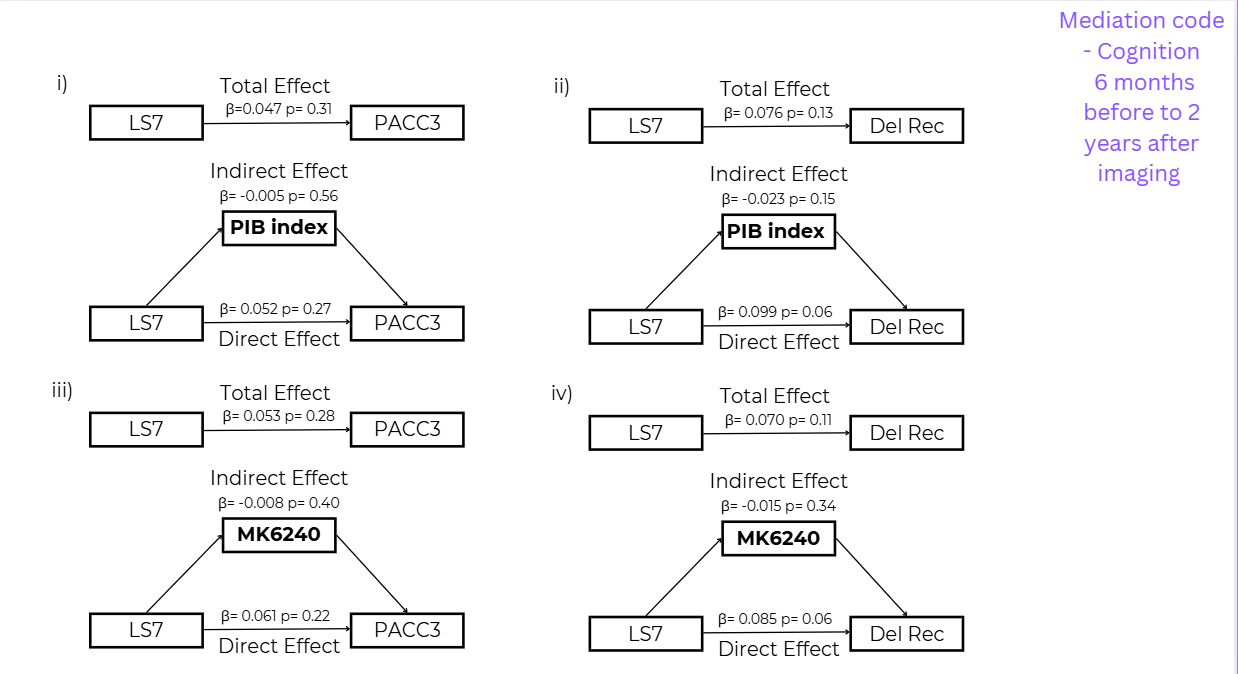 |

**Supplemental Figure 2: Mediation analysis of AD markers.** No significant indirect effects are seen for any of the AD markers (PIB index and MK6240) and cognition (PACC3 and Delayed Recall). Only the direct effect for LS7 on delayed recall was approaching significance. Mediation analysis results for the relationship between the Life Simple Seven (LS7) score, Alzheimer’s disease and cognitive outcomes. The panels show the total effect, direct effect, and indirect effect (via the mediator) of LS7 on cognitive composite scores, Pre-Clinical Alzheimer’s Cognitive Composite score 3 (PACC3; a and c) and delayed recall (Del Rec; b and d). Mediators include PIB index for amyloid (a, b; n=148) and MK6240 Mayo Temporal Composite for tau (c, d; n=147). Beta estimates (β) and p-values are provided for each pathway. Total effects reflect the overall association between LS7 and the cognitive composite score, indirect effects indicate the pathway through the mediator (Alzheimer’s disease marker), and direct effects show the relationship between LS7 and the outcome independent of the mediator. The results underscore the complexity of the relationships between cardiovascular health, cerebrovascular pathways, and cognitive performance.
